# Ring and community vaccination for Bundibugyo ebolavirus outbreak response: a stochastic network modelling study

**DOI:** 10.64898/2026.07.09.26357654

**Authors:** Jason R. Andrews, Placide K. Mbala, Patrick K. Mukadi, Jason Kindrachuk, Nicole A. Hoff, Anne W. Rimoin, Isaac I. Bogoch

## Abstract

**Background:** Vaccination with rVSV-ZEBOV is highly effective against Zaire ebolavirus, but protection against Bundibugyo ebolavirus (BDBV) is unknown. We used a stochastic network model of the 2026 Democratic Republic of the Congo BDBV outbreak to evaluate the potential impact of a partially cross-protective vaccine under operationally realistic conditions.

**Methods:** We developed a susceptible-exposed-infectious-recovered model on a two-layer household-community contact network calibrated to public data through July 5, 2026. The model incorporated stochastic detection, isolation, first- and second-degree contact tracing, reactive ring vaccination, and community vaccination. Base-case vaccine efficacy was 45% and included post-exposure protection against disease and mortality, with time to protection modelled as a continuous sigmoidal function. Sensitivity analyses varied vaccine efficacy, timing, case detection, and contact tracing.

**Findings:** Increasing case detection from 30% to 70% and contact tracing from 30% to 80% reduced deaths by 59·8% (IQR 54·6–65·0) compared with base operations without vaccination. Adding reactive ring vaccination reduced deaths by 65·5% (IQR 59·1–71·2) versus base, but by 13·6% (IQR −3·2 to 27·9) versus enhanced operations alone. Community vaccination at 20–80% coverage reduced deaths by 47·1–91·0%. With 50% community coverage, mortality reduction declined from 86% at outbreak declaration to 58% with a 14-day delay. Ring vaccination impact was sensitive to immune-onset timing, with incremental mortality benefit declining from 23% to 10% as the assumed immune-onset midpoint increased from 5 to 14 days.

**Interpretation:** Strengthening case finding and contact tracing is central to mortality reduction in BDBV outbreak response. If rVSV-ZEBOV provides clinically meaningful cross-protection against BDBV, reactive ring vaccination could provide a measurable but modest incremental benefit when operations are already strong. Larger mortality reductions require vaccination that reaches susceptible individuals before exposure, which is more consistent with rapid community vaccination in affected areas.

**Funding:** IIB is funded by the Canadian Institutes of Health Research (PJT-203753).

**Research in context:** *Evidence before this study:* We searched PubMed, medRxiv, WHO guidance, and outbreak-response reports as of July 6, 2026 for studies and reports of rVSV-ZEBOV vaccination, ring vaccination, Bundibugyo ebolavirus, Ebola virus disease transmission models, post-exposure vaccination, and vaccination during Democratic Republic of the Congo Ebola responses. Published evidence supports ring vaccination against Zaire ebolavirus, but there is limited direct evidence for rVSV-ZEBOV effectiveness against Bundibugyo ebolavirus, and limited quantitative evidence comparing reactive ring vaccination with broader community vaccination in the setting of delayed case detection and incomplete contact tracing.

*Added value of this study:* This study integrates contemporary 2026 Bundibugyo ebolavirus outbreak data with a stochastic household-community network model to quantify the separate and combined effects of case finding, contact tracing, reactive ring vaccination, community vaccination, timing, vaccine coverage, immune-onset kinetics, and behavioural risk compensation. Results demonstrate that strengthening case detection and contact tracing can avert a large fraction of mortality even without vaccination, and that reactive ring vaccination provides only modest additional benefit once these operations are strong. In contrast, rapid community vaccination in affected areas can produce substantially larger mortality reductions by reaching susceptible people before exposure, but its impact is highly sensitive to implementation delay, coverage, vaccine effectiveness, and behavioural assumptions.

*Implications of all the available evidence:* For Bundibugyo ebolavirus outbreaks, vaccination strategy should be framed as part of a layered response. Case finding and contact tracing remain essential for early detection, isolation, and quarantine, but the largest vaccine-attributable effects are expected when vaccination is deployed rapidly enough and broadly enough to create community immunity before exposure. Reactive ring vaccination remains operationally valuable but should not be assumed to substitute for timely community-level protection when cross-protection is incomplete.

## Introduction

The West African Ebola virus disease (EVD) epidemic of 2014–2016 transformed the global approach to outbreak control and accelerated the deployment of the recombinant vesicular stomatitis virus-Zaire (rVSV-ZEBOV) vaccine.^1,2^ The landmark Ebola Ça Suffit! trial conducted in Guinea demonstrated the effectiveness of ring vaccination, a strategy that targets contacts of individuals with EVD and contacts of contacts to establish a protective barrier around active transmission chains.^1,2^ Following these results, ring vaccination has been adopted as a central component of orthoebolavirus outbreak response and has since been deployed during multiple outbreaks in the Democratic Republic of the Congo (DRC) and elsewhere.^3–6^

Preclinical non-human primate studies provide biological evidence that rVSV-based vaccines can protect against BDBV challenge, and recent serological data found some evidence, albeit mixed, of cross-reactive BDBV glycoprotein responses after rVSVΔG-ZEBOV-GP vaccination in outbreak-affected populations in the DRC.^15–17^ These findings raise the question of the extent to which a partially cross-protective vaccine could reduce morbidity and mortality if deployed during a BDBV outbreak, in the context of real-world operational constraints.

Operational timing is especially important for ring vaccination. Cases must be detected, contacts elicited and traced, and vaccination delivered before exposure or early enough after exposure for protection to develop. In outbreaks with rapid transmission, delayed diagnosis, and incomplete tracing, a ring strategy may vaccinate many people after the biologically relevant window has passed. Broader community vaccination in affected areas could, in principle, shift more vaccine delivery before exposure, but would require substantially more doses and rapid deployment across affected communities. The relative effectiveness of these vaccine strategies with a partially protective vaccine, under realistic operational constraints, remains unclear.

To address this gap, we developed a stochastic network model calibrated to data from the ongoing BDBV outbreak in the Democratic Republic of the Congo. We compared enhanced case finding and contact tracing, reactive ring vaccination, community vaccination at different coverage levels, hybrid strategies, vaccination timing, immune-onset kinetics, and behavioural risk compensation. Our aim was not to forecast the trajectory of the ongoing epidemic, but to evaluate which combinations of surveillance, tracing, vaccination strategy, coverage, and timing could have the greatest impact on BDBV morbidity and mortality.

## Methods

### Study design and data sources

We developed a model of BDBV natural history and transmission to estimate the potential impact of alternative vaccination strategies using a partially cross-protective rVSV-ZEBOV-like vaccine during BDBV outbreaks. The analysis was designed to compare reactive ring vaccination, community vaccination in affected local populations, and hybrid strategies, and to evaluate the impact of operational assumptions rather than to produce a forecast of the 2026 outbreak. We therefore focused on relative effects across scenarios, including infections, deaths, and vaccine courses used as percentages of the simulated local transmission network.

Contemporary outbreak inputs were derived from public INRB-UMIE and partner data repositories for the 2026 DRC outbreak, including reported confirmed cases, confirmed deaths, and SitRep-derived surveillance indicators.^14,23^ Historical data from the 2007 Uganda BDBV outbreak and the 2012 Isiro, DRC BDBV outbreak were used to assess robustness of the results. We used these data to estimate the population-level epidemic trajectory and severity inputs, inform natural-history, transmission, and case-ascertainment assumptions, and compare whether qualitative conclusions were preserved under alternative conditions.

### Model structure and transmission dynamics

We developed a stochastic susceptible-exposed-infectious-recovered (SEIR) model over a two-layer contact network designed to capture the local structure of BDBV transmission. The first layer represented household and caregiving exposure through fully connected clusters with mean size approximately five. The second layer represented community exposure through a negative-binomial degree distribution, allowing heterogeneity in the number of contacts. This structure was intended to capture both intense within-household exposure and occasional high-degree community transmission events, which are characteristic concerns in filovirus outbreaks.

### Parameterisation and calibration

Baseline natural-history assumptions were chosen from BDBV-specific and Ebola virus disease literature and then updated with contemporary outbreak data where available. The mean incubation period was 8·5 days and the mean infectious period was 6·0 days. The unvaccinated case fatality risk was 45·4%, based on an adjusted estimate that accounted for the lag between case confirmation and clinical outcome ^24^; the latest naive cumulative case fatality in public SitRep-derived data was 32·1%.

To parameterise time-varying transmission, we estimated Rt from daily case notifications using a renewal-equation framework ^25^ and a generation-interval distribution parameterised from Ebola virus disease serial-interval estimates.^26^ Because comprehensive individual-level contact-tracing data were not available, we did not attempt to reconstruct the exact real-world contact network.

Instead, we used a representative synthetic network and calibrated the model to the population-level epidemic trajectory by scaling the per-edge transmission hazard according to the estimated Rt trajectory. Case detection was represented as a reporting probability informed by public surveillance indicators and varied across scenario values.

Broad exploratory grids used smaller local networks to examine large parameter spaces. Final estimates used 100,000-person local networks and high-replicate stochastic simulation ensembles for the main manuscript scenarios. Population-size sensitivity analyses were interpreted as robustness checks for local transmission-network scale rather than forecasts of the total population at risk across DRC health zones.

#### Vaccination strategies and operational constraints

Reactive ring vaccination was triggered by stochastic case detection. After detection, direct contacts could be vaccinated according to tracing and uptake assumptions. Extended ring vaccination added contacts of contacts (Ring 2) with an additional delay and lower coverage. Vaccinated individuals remained eligible for monitoring and case detection in the final model implementation, so vaccination did not remove contacts from surveillance.

Community vaccination was modelled as vaccination of a specified fraction of the local network in affected communities. Coverage scenarios ranged from 20% to 80% for primary strategy comparisons and 10% to 80% for contour analyses. Hybrid strategies combined 20–80% community vaccination coverage with reactive ring vaccination. Timing scenarios evaluated 50% community coverage beginning at outbreak declaration, 7 days after declaration, or 14 days after declaration.

Because the first laboratory-confirmed case and outbreak declaration occurred on May 15, 2026, model day 0 should be interpreted as this date. The day-0 community vaccination scenario represents an aggressive campaign beginning at declaration, modeled as a linear rollout achieving target coverage over 14 days, rather than pre-existing population immunity before recognition of the outbreak. Sensitivity analyses varied the initiation of this 14-day rollout to 7 and 14 days post-declaration. We also evaluated hybrid strategies, where community vaccination and ring vaccination were implemented jointly, to test the incremental benefit of ring vaccination once community vaccination had already been deployed.

### Vaccine efficacy, immune onset, and therapeutic rescue

The base vaccine efficacy assumption was 45%, representing a conservative partial cross-protection scenario approximately half the reported efficacy of rVSV-ZEBOV against Zaire ebolavirus in clinical trials and observational studies. To avoid biologically implausible combinations, this value was applied consistently across vaccine-mediated protection against infection, post-exposure benefit, and mortality reduction among vaccinated infected individuals. Sensitivity analyses varied the vaccine-effectiveness parameter and scaled these vaccine effects together.

Vaccine-mediated protection was delayed after vaccination. In base analyses, protection accrued over a 10-day immune-onset window; sensitivity analyses explored alternative timing assumptions. For vaccinated individuals who nevertheless became infected, the vaccine could reduce mortality only when delivered early enough relative to illness progression.

For sensitivity analyses, we varied vaccine-effect assumptions, timing of vaccine deployment, dose-use constraints, behavioural risk compensation, and historical transmission parameterisations. These analyses were designed to test whether the qualitative ordering of strategies was robust to uncertainty in biological cross-protection and operational implementation.

### Scenarios and outcomes

Primary outcomes were cumulative infections, deaths, and vaccine courses administered. Results are reported as percent reductions relative to the explicit comparator shown for each analysis.

Strategy comparisons include reductions versus base operations, reductions versus enhanced operations, and incremental vaccine-attributable reductions when added to enhanced case finding and contact tracing.

Base operations assumed 30% index-case detection and 30% contact-tracing coverage, representing lower-intensity response conditions with incomplete case ascertainment and limited contact follow-up similar to observations in some conflict-affected DRC outbreaks.^27,28^ Enhanced operations assumed 70% index-case detection and 80% contact-tracing coverage, representing strengthened case finding, contact investigation, and isolation capacity during a scaled outbreak response. The main manuscript scenarios were: base operations without vaccination; enhanced operations without vaccination; enhanced operations plus reactive ring vaccination; community vaccination at 20%, 40%, 60%, and 80% coverage. Main results reflect 100,000-person local transmission networks and are reported as relative reductions in cumulative infections, deaths, or vaccine courses administered compared with a prespecified comparator.

The primary estimate was the relative reduction in cumulative deaths under alternative operations and vaccination strategies. Absolute percentages of the simulated local network were used internally to compare stochastic trajectories on a common scale, but the manuscript emphasises relative effects because the simulated network represents a local transmission environment rather than the total population at risk in the DRC outbreak.

### Behavioural risk compensation

We evaluated the potential impacts of behavioural risk compensation as an exploratory scenario. In these scenarios, vaccinated individuals could have increased effective contact rates, reflecting a condition in which perceived protection led to relaxation of infection-control behaviours. These analyses tested whether incomplete cross-protection could be offset or undermined by behavioural adaptation, modelled as increased effective contact rates across a range of values.

### Sensitivity analyses

We conducted sensitivity analyses for natural-history parameters, historical transmission settings, vaccine effectiveness, immune-onset assumptions, community vaccination coverage and timing, behavioural risk compensation, dose use, network population size, case detection and contact tracing. For immune onset, we varied the midpoint of a continuous vaccine-mediated protection profile at 5, 10, and 14 days after vaccination, representing optimistic early protection, the 10-day protection window used in rVSV-ZEBOV ring-vaccination efficacy analyses, and slower onset consistent with detectable humoral responses around 10–14 days. Operational ring-vaccination analyses varied index-case detection and contact coverage for Ring 1 and Ring 2 vaccination, using matched no-vaccination comparators with the same detection and tracing assumptions. We also examined how delays in community vaccination after outbreak declaration affected mortality reduction, and how vaccine courses used translated into incremental benefit across ring, community, and hybrid strategies.

### Model reporting and reproducibility

Key model assumptions are summarised in Table 1, with additional implementation details reported in the appendix. Code, input data, generated parameter files, simulation outputs, and figure-generation code are publicly available at https://github.com/jasonandr/bundibugyo-ebov-vaccination.

**Table 1:** Key model parameters, ranges and data sources.

| Parameter | Baseline or range | Source |
| --- | --- | --- |
| Incubation period, mean | 8.5 days | [28,29] |
| Infectious period, mean | 6.0 days | [28,29] |
| Unvaccinated case fatality risk | 45.4% | Estimated from public 2026 data [14,23,27] using a delay-adjusted severity approach [24]; historical comparison [28,29] |
| Vaccine effectiveness against BDBV (VE) | 45% in base case; varied in sensitivity analyses | Assumed scenario value, applied consistently to protection against infection, post-exposure benefit, and mortality reduction |
| Immune-onset window | 10-day base immune-onset window; alternative timing in sensitivity analyses | Assumed; informed by rVSV-ZEBOV efficacy window and immunogenicity data [1-3] |
| Post-exposure mortality reduction | Scaled with the overall vaccine-effect assumption | Assumed; scenario analysis |
| Case detection probability | 30% in base operations; 70% in enhanced operations and main vaccine comparisons | Scenario values informed by 2026 surveillance data [14,23] |
| Detection delay | 4 days from symptom onset | Assumed |
| Reactive ring vaccination | Direct contacts; extended ring includes contacts of contacts with additional tracing delay | Assumed; intervention comparison [1,2,4] |
| Community vaccination coverage | 20%, 40%, 60%, and 80% in primary comparisons; broader range in contours | Scenario analyses |
| Community vaccination timing | At outbreak declaration, declaration +7 days, and declaration +14 days | Scenario analyses |

### Ethics and data governance

The analysis used aggregated, publicly available outbreak reports and de-identified historical data sources. No individual-level identifiable patient data were used in this modelling analysis. The work should therefore be considered secondary analysis of public health surveillance data rather than human-subjects research requiring participant consent.

## Results

The latest public data available for this analysis reported 1624 confirmed cases and 521 confirmed deaths in the DRC by July 5, 2026. The corresponding crude confirmed case fatality risk was 32·1%; after accounting for the lag from case confirmation to outcome, the adjusted case fatality risk used in baseline simulations was 45·4%. The estimated Rt ranged from 1·07 to 6·06 across the study period and was 1·13 at the end of the series. The peak estimated Rt was similar in magnitude to historical reconstructions for the 2007 Bundibugyo, Uganda, and 2012 Isiro, DRC BDBV outbreaks, which had maximum estimated Rt values of 2·37 and 1·33, respectively.

Enhanced operations alone had a large effect before vaccination was added. Compared with base operations without vaccination, enhanced case finding, contact tracing, and isolation reduced median infections by 60·2% (IQR 55·0–65·0) and median deaths by 59·8% (IQR 54·6–65·0).

Reactive ring vaccination produced modest incremental reductions when added to enhanced operations (Figure 2). Enhanced operations plus reactive ring vaccination reduced median deaths by 65·5% (IQR 59·1–71·2) compared with base operations, but the incremental mortality reduction compared with enhanced operations alone was 13·6% (IQR −3·2 to 27·9). The corresponding incremental infection reduction was 10·9% (IQR −6·1 to 25·5), indicating that under the base vaccine-effect assumptions the additional effect of reactive ring vaccination was modest once case finding and contact tracing were already strong.

**Figure 1.**
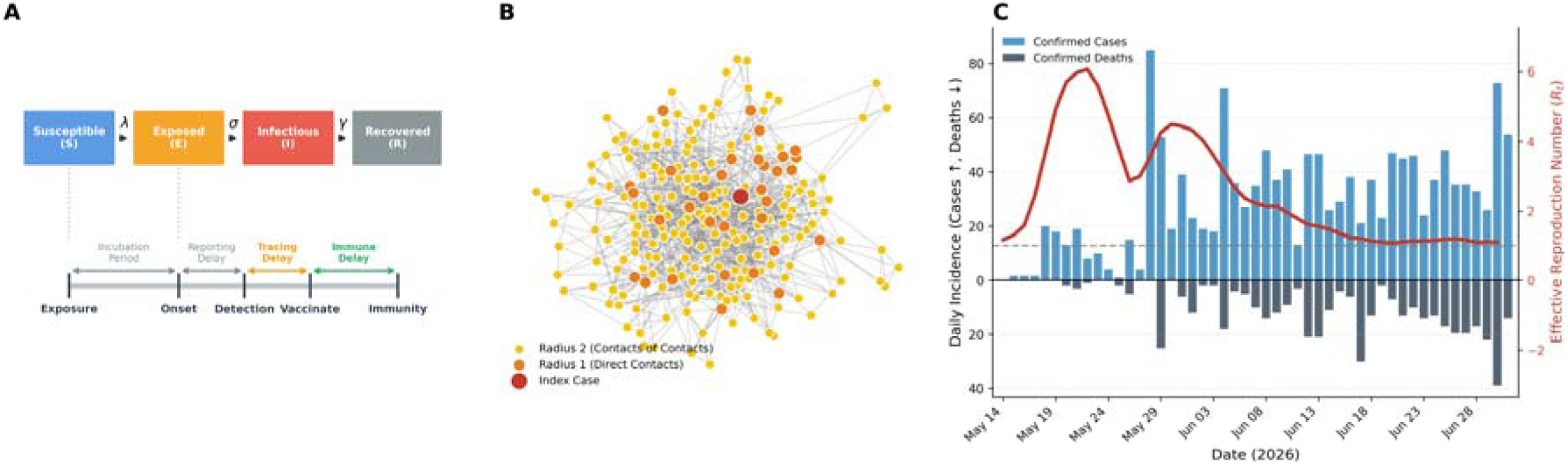
Model structure and outbreak calibration. (A) Natural-history model of Bundibugyo virus disease and the operational timeline for detection, isolation, contact tracing, and vaccination. (B) Two-layer household-community contact network underlying the stochastic transmission model. (C) Estimated effective reproduction number (Rt) alongside daily confirmed cases from the 2026 Bundibugyo ebolavirus outbreak.

**Figure 2.**
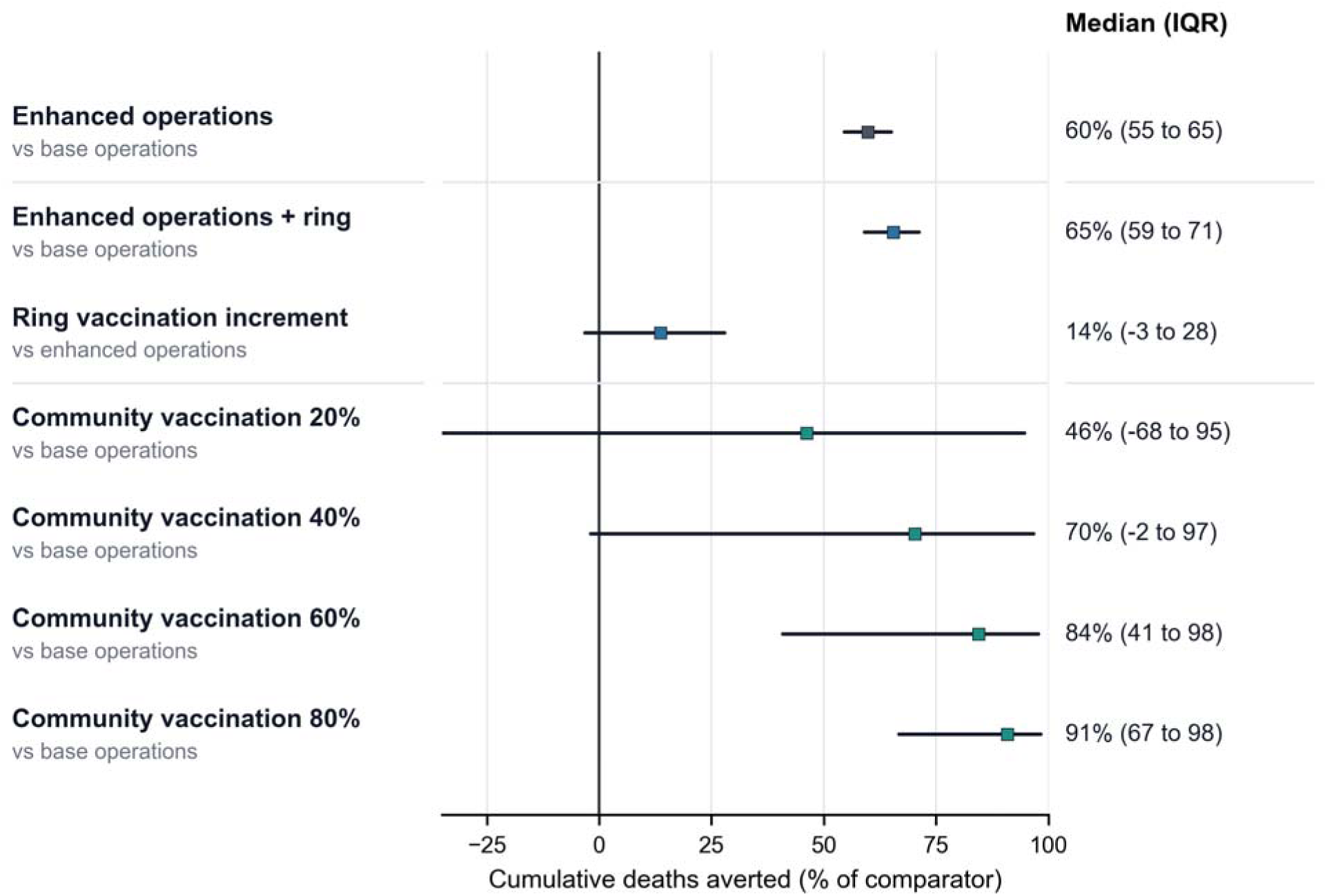
Impact of public health and vaccination strategies under base vaccine efficacy assumptions. The forest plot shows median cumulative deaths averted and interquartile ranges across paired stochastic simulations. Comparators are shown in the row labels: enhanced operations and enhanced operations plus ring vaccination are shown versus base operations; the ring-vaccination increment is shown versus enhanced operations; community vaccination scenarios are shown versus base operations. Vaccine scenarios assume a base vaccine efficacy of 45%, applied consistently to protection against infection, post-exposure benefit, and mortality reduction.

Community vaccination produced a larger and more consistent impact. At 20%, 40%, 60%, and 80% coverage, median mortality reductions versus base operations were 46·2% (IQR −68·2 to 94·7), 70·3% (IQR 0·0 to 96·9), 84·4% (IQR 40·5 to 97·9), and 90·9% (IQR 66·8 to 98·4), respectively. The corresponding median infection reductions were 44·2%, 68·3%, 82·0%, and 88·3%. Figure 3 trajectories showed the same qualitative pattern. By day 80, median cumulative mortality reduction compared with base operations was 60% (IQR 55–65) for enhanced operations, 65% (IQR 59–71) for enhanced operations plus ring vaccination, and 71% (IQR 0–97) for 40% community vaccination.

**Figure 3.**
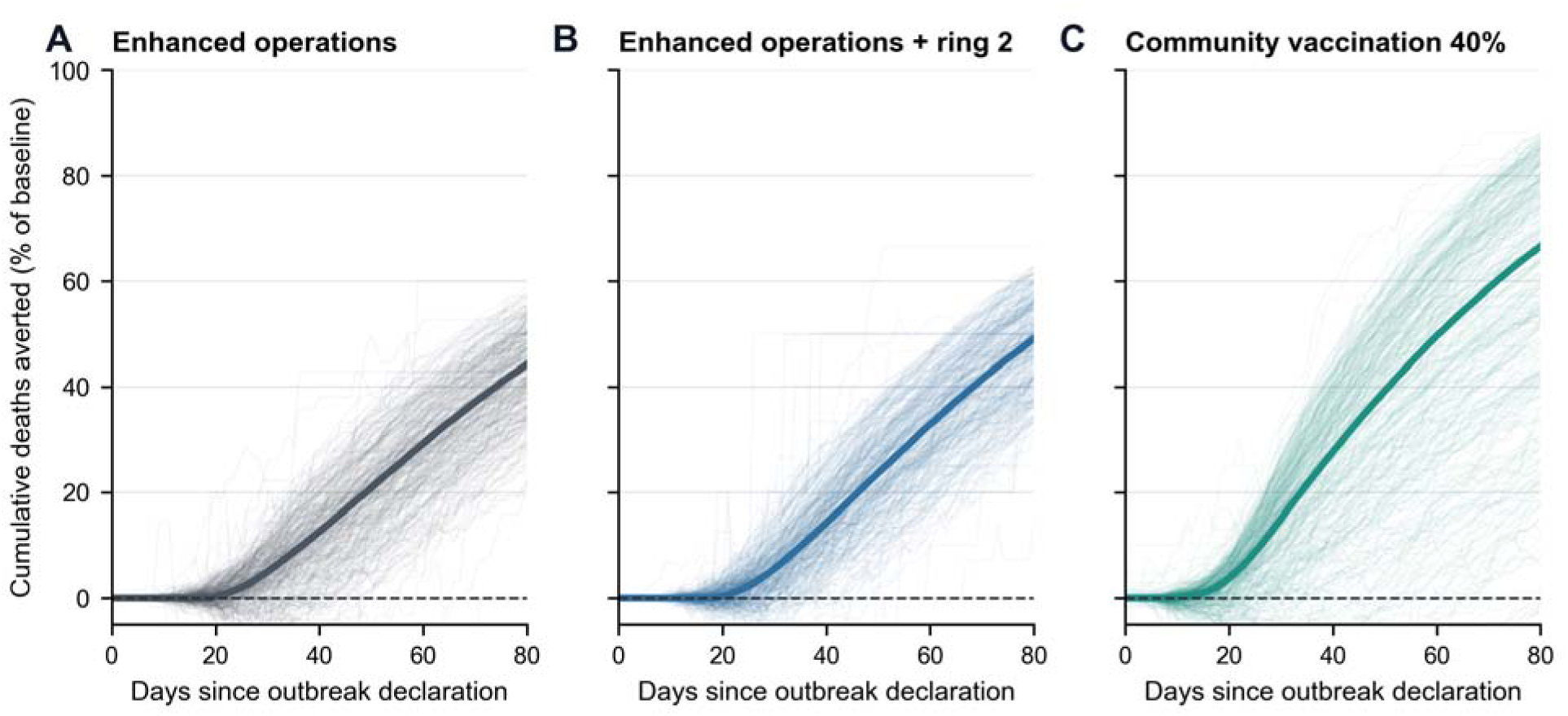
Dynamic mortality trajectories by response strategy. Lines show stochastic replicate trajectories within the final-outcome interquartile range and thick lines show median cumulative deaths averted as a percentage of the base-operations baseline. (A) Enhanced operations. (B) Enhanced operations plus ring 2 vaccination. (C) Community vaccination at 40% coverage. Time is measured from outbreak declaration on May 15, 2026; trajectories are shown through day 80.

Contour analysis showed that index case detection and contact tracing coverage jointly determined the benefit of enhanced operations relative to base operations (Figure 4A). The base-operations point at 30% index-case detection and 30% contact-tracing coverage lay near the low-benefit region, whereas higher detection and tracing moved the response into substantially larger mortality reductions. Furthermore, the analysis demonstrates that if operational capacities fall below the base-case levels of 30% detection and tracing, outbreak morbidity and mortality will worsen substantially compared to the baseline trajectory.

**Figure 4.**
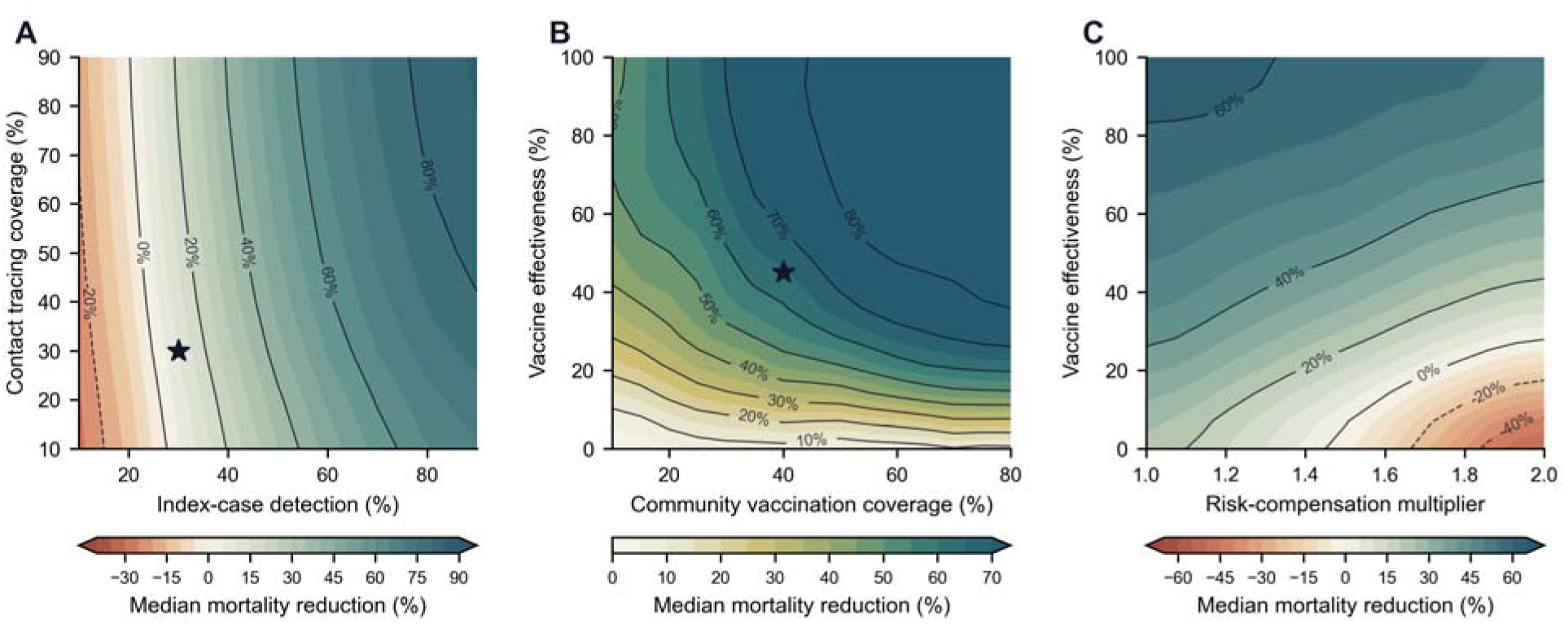
Operational and vaccine-effect contours for mortality reduction. (A) Median mortality reduction for enhanced operations versus base operations across index-case detection and contact-tracing coverage. The star marks the base-operations assumption of 30% detection and 30% contact-tracing coverage; negative values indicate settings worse than the paired base comparator. (B) Median mortality reduction across community vaccination coverage and vaccine effectiveness; the star marks 40% community coverage and 45% vaccine effectiveness. (C) Median mortality reduction under behavioural risk-compensation multipliers and vaccine effectiveness. Values below 0% indicate scenarios in which increased effective contact rates offset vaccine benefit.

Community-vaccination contour analyses showed that impact was jointly determined by coverage and vaccine effectiveness (Figure 4B). Under the base assumption of 45% vaccine effectiveness, increasing community coverage moved scenarios across a steep gradient of mortality reduction. At lower vaccine efficacy values, higher coverage remained beneficial but produced smaller gains, while at higher vaccine efficacy values, moderate-to-high coverage produced large reductions.

Behavioural risk-compensation analyses suggested that increased effective contact rates after vaccination could erode vaccine benefit when vaccine effect was low (Figure 4C). At the base 45% vaccine efficacy assumption, the model retained mortality reductions under modest risk compensation, but the margin narrowed as the risk-compensation multiplier increased and vaccine efficacy decreased. These scenarios should be interpreted as stress tests of behavioural and infection-control assumptions, not as evidence that risk compensation will occur during BDBV outbreak response.

Timing of vaccine introduction strongly modified the expected benefit of community vaccination (Figure 5A). With 50% community coverage, median mortality reduction was 86% (IQR 76–91) when vaccination began at outbreak declaration, 67% (IQR 54–77) with a 7-day delay after declaration, and 58% (IQR 42–70) with a 14-day delay after declaration. Because model day 0 corresponds to May 15, 2026, this scenario is best interpreted as a rapid campaign at declaration rather than pre-existing immunity before recognition of the outbreak.

**Figure 5.**
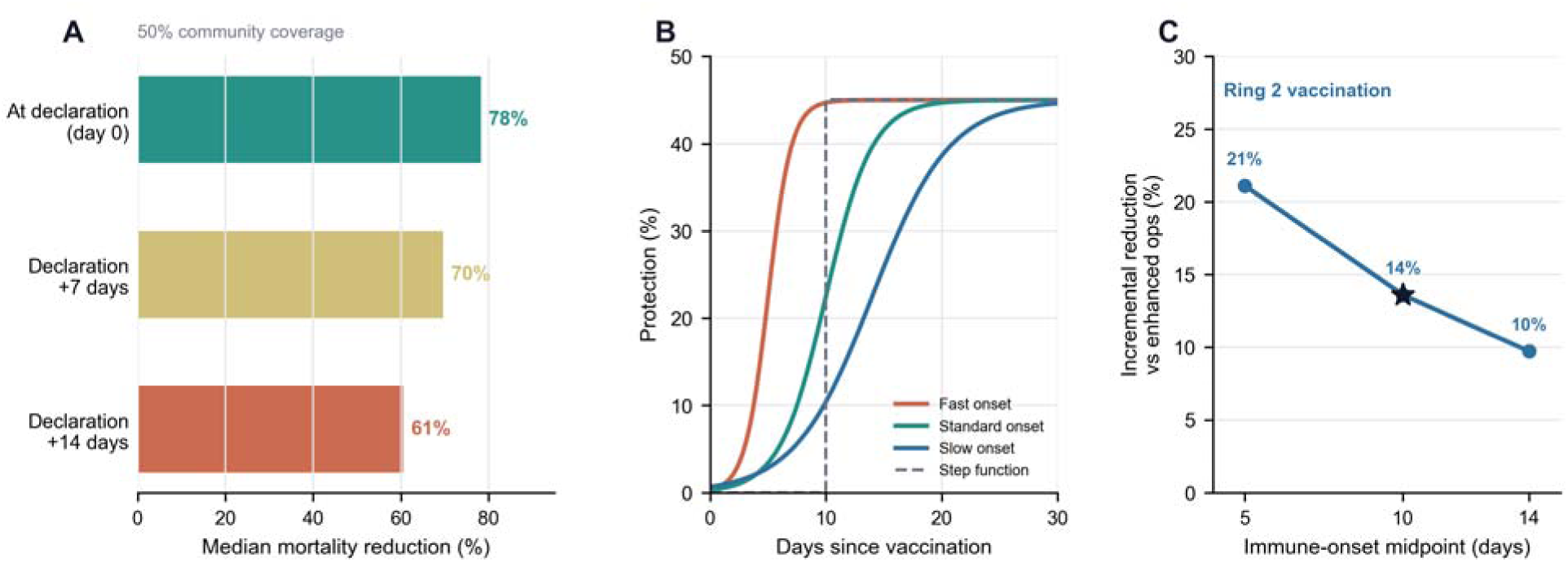
Timing of vaccine delivery and immune onset. (A) Median mortality reduction with 50% community vaccination coverage when vaccination begins at outbreak declaration or 7 or 14 days after declaration. (B) Sigmoidal immune-onset profiles used to represent fast, standard, and slow development of vaccine-mediated protection, with a step-function comparator. (C) Incremental mortality reduction from ring 2 vaccination versus enhanced operations without vaccination under alternative immune-onset midpoint assumptions.

Immune-onset timing also impacted the incremental benefit of reactive ring vaccination (Figure 5B-C). The model represented protection as continuous sigmoidal onset profiles, with midpoint values at 5, 10, and 14 days after vaccination. Shifting the immune-onset midpoint from 5 to 10 to 14 days reduced the incremental mortality reduction of ring 2 vaccination from 23% (IQR 5–37) to 15% (IQR −3 to 29) and 10% (IQR −11 to 26), respectively, relative to enhanced operations without vaccination.

## Discussion

Bundibugyo virus is an emerging orthoebolavirus that has caused several outbreaks in central Africa, including the ongoing outbreak in the Democratic Republic of the Congo. In this modelling study, we evaluated how a partially cross-protective rVSV-ZEBOV-like vaccine might perform under operational conditions relevant to Bundibugyo virus disease response. Relative to enhanced case finding and contact tracing without vaccination, reactive ring vaccination produced modest incremental reductions in infections and deaths, whereas rapid community vaccination produced larger and more consistent reductions when sufficient coverage was achieved. The most consistent signal across analyses was that population-level benefit depended strongly on whether vaccination reached susceptible individuals before exposure and whether vaccine-mediated protection developed quickly enough to affect outcomes.

Vaccination has transformed outbreak response for Zaire ebolavirus. The rVSV-ZEBOV vaccine has shown high efficacy and is now central to Ebola virus disease control,^1–6^ but no licensed vaccine is currently available for Bundibugyo virus disease. Preclinical studies suggest that

VSV-based vaccines can provide some heterologous protection against BDBV, and serological studies from outbreak-affected populations provide supportive evidence of cross-reactive responses.^15–17^ The policy question is therefore not simply whether a vaccine has biological activity against BDBV, but whether that activity can be translated into population-level benefit under the time constraints of outbreak response.

Our results confirm that biological vaccine properties matter, but find that the magnitude of their effects at the population level are strongly determined by the performance of the public health system in which they are deployed. Higher VE and broader coverage improved outcomes, as expected, but the difference between reactive ring vaccination and community vaccination was driven largely by timing. Ring vaccination is triggered by detection of an infectious person; if detection, contact elicitation, tracing, and vaccination occur after many contacts have already been exposed, the ring can still support surveillance and may provide post-exposure benefit, but its ability to prevent onward transmission is limited. Community vaccination uses more doses, but it shifts protection earlier in the transmission and natural history process.

One concern with deploying a partially protective vaccine against a pathogen with high lethality is behavioural risk compensation, whereby vaccinated individuals might reduce adherence to other infection-control practices because they perceive themselves to be protected.^20^ This could include delaying care-seeking, providing direct care to family members or community members with BDBV disease, or participating in unsafe burial practices. Our exploratory analyses should not be interpreted as evidence that such behaviour will occur during BDBV outbreaks. However, they show that when vaccine cross-protection is incomplete, increased contact rates after vaccination could offset part of the prophylactic benefit. Risk communication should therefore present vaccination as one component of outbreak control, alongside rapid isolation, safe and dignified burials, timely care seeking, contact tracing, and sustained community engagement.^21,22^

These findings have implications for both vaccine evaluation and outbreak response for BDBV or other related pathogens. If a cross-reactive vaccine were deployed during a BDBV outbreak, its observed public-health effect would depend not only on the product’s biological efficacy but also on the response system in which it was used. Low ascertainment or delayed tracing could make a biologically active vaccine appear ineffective at population level, whereas rapid detection and efficient early deployment could allow even partial protection to avert deaths. Modelling studies and field evaluations should therefore report operational assumptions as carefully as biological efficacy assumptions.

The results of this study should be interpreted within the context of several limitations. Vaccine efficacy against BDBV is unknown, and the VE and post-exposure mortality-reduction values evaluated here should be interpreted as scenario assumptions rather than estimates for a licensed product. We used simulated contact networks rather than observed contact-tracing data from the 2026 outbreak. The scenario grids used fixed transmission assumptions and did not propagate full uncertainty in time-varying reproduction numbers, case detection, tracing delays, vaccine acceptance, or behavioural responses. Although we used 100,000-person networks, larger health zones and urban transmission settings could involve more spatial structure, mobility, and heterogeneity than represented here. The results are therefore most useful for identifying operational dependencies and plausible thresholds, rather than forecasting anticipated case counts or deaths.

We did not make true pre-outbreak preventive vaccination a main scenario. Preventive population vaccination is not routinely implemented for ebolaviruses, and if a population had substantial immunity before recognition of a subcritical outbreak, the model would predict large or complete prevention by construction. The more policy-relevant question for outbreaks is how much benefit could be achieved by vaccination beginning at or shortly after declaration in affected communities.

As ebolavirus outbreaks continue to occur in settings with fragile health systems, population mobility, and substantial operational constraints, the effectiveness of vaccination will depend on both biological and implementation realities. A partially cross-protective vaccine could reduce morbidity and mortality, but its value will be determined principally by how quickly cases are detected, how completely contacts and affected communities are reached, and how effectively communities remain engaged. Understanding where reactive rings fail, and when broader targeted vaccination is feasible, is essential for designing response strategies that translate vaccine activity into durable epidemic control.

## Contributors

JRA and IIB conceived the idea and conducted the modelling. All authors contributed to the interpretation of data, writing, and editing of the manuscript.

## Data Availability

All data produced in the present work are contained in the manuscript

