## Supplementary material for "Ring and community vaccination for Bundibugyo ebolavirus outbreak response: a stochastic network modelling study": Supplument

**Table of Contents**

**1. Supplementary Methods**

Model overview and event scheduling

Contact network

Transmission process and use of Rt

Case detection, monitoring, and isolation

Ring tracing, queueing, and vaccination scheduling

Community vaccination

Vaccine effects and immune onset

Outcomes and stochastic summaries

**2. Supplementary Results**

**3. Supplementary Figures**

**Supplementary Table S1. Final high-replicate scenario estimates**

Supplementary Figure S1. Historical 2007 Bundibugyo dynamics

Supplementary Figure S2. Historical 2012 Isiro dynamics

Supplementary Figure S3. Dose-use frontier

Supplementary Figure S4. Delivery-window analysis

### 1. Supplementary Methods

#### Model overview and event scheduling

We used a stochastic, individual-based SEIR model implemented on a fixed contact network. Individuals occupied one of six operational states: susceptible, exposed, infectious, recovered, vaccinated susceptible, or isolated infectious. The simulation used a continuous-time event queue. Events included exposure, symptom onset, case detection, contact vaccination, recovery, and community vaccination. Event times were processed in chronological order using a priority queue, which allowed infection, isolation, tracing, and vaccination delays to interact within each stochastic realization.

Exposed individuals progressed to symptom onset after an exponentially distributed latent period with mean 8.5 days. Infectious individuals recovered after an exponentially distributed infectious period with mean 6.0 days unless detected and isolated first. Isolation did not retroactively remove infections that had already occurred, but it blocked future scheduled exposure attempts from that source. Death was assigned at symptom onset using the scenario-specific case fatality risk, modified by vaccination status and time since vaccination as described below.

#### Contact network

The contact network was generated as a two-layer graph. The first layer partitioned individuals into household or caregiving clusters, with cluster sizes drawn as 1 + Poisson(mean household size – 1), ensuring a minimum cluster size of one, and all members within a cluster connected to one another. The second layer added community contacts from an overdispersed negative-binomial degree distribution. For the main simulations the mean community degree was 5 and the variance was 25, creating a heavy-tailed degree distribution intended to represent heterogeneity in community exposure opportunity. Final manuscript estimates used 100,000-person local transmission networks, with 10,000 stochastic replicates for the principal reactive-ring and community-vaccination scenario summaries.

The model uses this network as a local transmission environment rather than as a census of the total outbreak population. Consequently, results are reported primarily as relative reductions in infections, deaths, or vaccine courses compared with explicit within-model comparators. The household layer increases local clustering and repeated exposure opportunities, whereas the community layer creates overdispersed contact opportunity.

#### Transmission process and use of Rt

Transmission was represented as a thinned continuous-time contact process. When an individual became infectious, the model scheduled candidate exposure events to susceptible or vaccinated neighbours. Candidate event times were drawn from an exponential distribution with rate tau_max. At candidate time t, the event was accepted with probability Rt(t) / Rmax, where Rt(t) is the daily effective reproduction-number trajectory used for calibration and Rmax is the maximum value of that trajectory. Thus the effective per-edge transmission hazard was proportional to tau_max x Rt(t) / Rmax.

For each accepted candidate exposure, vaccine-mediated protection was evaluated before infection occurred. If a vaccinated susceptible individual was protected at that time, the event was discarded and the individual remained susceptible or vaccinated susceptible. If not protected, the individual entered the exposed state and an onset event was scheduled. This implementation allowed vaccination to affect infection risk before exposure while preserving stochastic variation in who was reached before infection.

The Rt trajectories used in the main and historical robustness analyses were estimated from outbreak incidence data and then used to scale the daily transmission hazard in the network model. This calibration approach separates the shape of the time-varying epidemic trajectory from the stochastic contact process: the network generates individual-level infection, tracing, and vaccination events, while the Rt array imposes the population-level temporal forcing used for each outbreak scenario.

#### Case detection, monitoring, and isolation

At symptom onset, an infectious case was detected with the scenario-specific detection probability. Detection occurred after the detection delay unless the individual was already under monitoring from prior contact tracing. Monitored individuals used a shortened one-day detection delay, representing active follow-up of traced contacts. Detection before recovery moved an infectious individual to the isolated state and initiated ring tracing and vaccination if the scenario allowed vaccination at that time.

Base operations and enhanced operations differed through the reporting and contact-tracing assumptions. In the base-operations comparator, both index-case detection and tracing coverage were lower. Enhanced operations increased case detection and contact tracing before vaccination was added, allowing the analyses to separate benefits attributable to surveillance and isolation from incremental vaccine-attributable benefit.

#### Ring tracing, queueing, and vaccination scheduling

When an infectious case was detected, the model identified ring contacts by search on the contact network. Radius 1 included direct contacts of the detected case; radius 2 additionally included contacts of contacts. For each eligible contact, tracing occurred with probability equal to the scenario-specific tracing coverage. Traced contacts were marked as monitored regardless of whether they subsequently accepted vaccination, so traced vaccinated contacts remained eligible for enhanced case detection if they later developed disease.

Vaccination was then scheduled only for traced contacts who accepted vaccine and had not already received vaccine. Direct contacts used the base tracing delay. Radius 2 contacts used the base tracing delay plus an additional delay term, reflecting the extra time required to identify and reach contacts of contacts. The default extended-ring uptake multiplier reduced radius 2 uptake relative to radius 1.

#### Community vaccination

Community vaccination was implemented as a time-limited campaign beginning at outbreak declaration or after a scenario-specific delay. For a target coverage level c, the model randomly selected approximately cN individuals from the local transmission network. Vaccination was then distributed evenly across the rollout period, with a base-case duration of 14 days. Each day, the model vaccinated a daily quota equal to the target number divided by the rollout duration.

Community vaccination could reach susceptible and exposed individuals. Susceptible vaccinated individuals entered the vaccinated susceptible state; exposed vaccinated individuals remained exposed but retained their vaccination time, allowing post-exposure effects on disease progression and mortality to be evaluated. Individuals already scheduled through a ring were not double-counted when community vaccination occurred first.

#### Vaccine effects and immune onset

The base vaccine-effect parameter was applied consistently to protection against infection and to reduction in mortality risk among vaccinated infected individuals. In the base case analyses, protection followed a sigmoid function VE(d) = VEmax / [1 + exp{-k(d - d0)}], where d is days since vaccination, VEmax is the maximum vaccine effect, k controls the steepness of onset, and d0 is the midpoint of immune onset. We also compared this with a binary immune delay, where infection protection was zero before the immune delay and equal to the scenario efficacy after the delay.

Mortality protection used the same time-since-vaccination structure but scaled the transition from the unvaccinated case fatality risk to the vaccinated case fatality risk. At symptom onset, the model computed CFR(d) = CFR0 - B(d) x (CFR0 - CFRv), where CFR0 is the unvaccinated case fatality risk, CFRv is the vaccinated case fatality risk after full effect, and B(d) is the time-dependent therapeutic benefit function. This allowed vaccination received after infection but before symptom onset to reduce mortality without necessarily preventing infection.

#### Outcomes and stochastic summaries

For each stochastic replicate, the model recorded cumulative infections, cumulative deaths, and vaccine courses administered. Scenario effects were calculated relative to the explicit comparator for each analysis: enhanced operations and community vaccination were compared with base operations where indicated, whereas incremental reactive ring-vaccination effects were compared with enhanced operations without vaccination.

Reported uncertainty intervals are empirical interquartile ranges across stochastic simulations under fixed scenario assumptions. They quantify stochastic variation in outbreak realizations and operational pathways, not full parameter uncertainty. Paired comparisons used shared simulation indices where available so that intervention and comparator runs were aligned by stochastic replicate structure as closely as possible.

### 2. Supplementary Results

#### Historical robustness analyses

To evaluate whether the primary findings were sensitive to baseline transmission assumptions, we conducted robustness analyses using empirical estimates derived from earlier BDBV outbreaks. Historical incidence reconstructions from the 2007 Bundibugyo outbreak and the 2012 Isiro outbreak were used to estimate effective reproduction-number trajectories. We recalibrated the stochastic network transmission hazard to these historical dynamics and evaluate the impact of vaccination strategies.

#### Final high-replicate scenario estimates

Final estimates used 100,000-person local transmission networks. Table S1 reports median infection reductions, mortality reductions and vaccine courses. The strongest mortality reductions occurred when vaccination reached susceptible individuals before exposure, particularly in rapid community-vaccination scenarios. Reactive ring vaccination provided a measurable but smaller incremental benefit when added to enhanced case finding and contact tracing.

#### Supplementary Table S1. Final high-replicate scenario estimates

Values are medians across stochastic simulations. Mortality reductions are relative to the comparator stated in the row.

| **Scenario** | **Median infection reduction** | **Median mortality reduction** | **Mean vaccine courses (%)** | **Interpretation** |
| --- | --- | --- | --- | --- |
| Base operations, no vaccination | Reference | Reference | 0.0 | Comparator for operational improvement |
| Enhanced operations, no vaccination | 61% vs base | 61% vs base | 0.0 | Case finding, tracing, and isolation before vaccination |
| Enhanced operations + reactive ring vaccination | 10% vs enhanced; 65% vs base | 13% vs enhanced; 66% vs base | 8.1 | Incremental vaccine effect after operational strengthening |
| Community vaccination, 20% coverage | 75% vs base | 76% vs base | 20.0 | Coverage-response analysis |
| Community vaccination, 40% coverage | 83% vs base | 85% vs base | 40.0 | Base community coverage in main comparisons |
| Community vaccination, 60% coverage | 89% vs base | 90% vs base | 60.0 | Coverage-response analysis |
| Community vaccination, 80% coverage | 92% vs base | 94% vs base | 80.0 | High-coverage scenario |
| 50% community vaccination at declaration | 87% vs base | 88% vs base | 50.0 | Timing analysis |
| 50% community vaccination, declaration +7 days | 81% vs base | 83% vs base | 49.9 | Timing analysis |
| 50% community vaccination, declaration +14 days | 76% vs base | 78% vs base | 49.8 | Timing analysis |
| Reactive ring vaccination, 5-day immune-onset midpoint | 16% vs enhanced | 20% vs enhanced | 7.5 | Immune-onset sensitivity |
| Reactive ring vaccination, 14-day immune-onset midpoint | 7% vs enhanced | 9% vs enhanced | 8.4 | Immune-onset sensitivity |

#### Supplementary Table S2. Historical outbreak robustness estimates

| Scenario | Outbreak | Median infection reduction (%) | Median mortality reduction (%) |
| --- | --- | --- | --- |
| Enhanced Operations (vs Base) | 2007 | 2.5% | 2.6% |
| Reactive Ring Vaccination (vs Enhanced) | 2007 | 1.8% | 2.7% |
| Community Vaccination (vs Base) | 2007 | 32.5% | 43.5% |
| Enhanced Operations (vs Base) | 2012 | 3.7% | 3.7% |
| Reactive Ring Vaccination (vs Enhanced) | 2012 | 2.0% | 2.8% |
| Community Vaccination (vs Base) | 2012 | 36.6% | 47.0% |

### Values are medians across stochastic simulations using historical transmission profiles.

### 3. Supplementary Figures

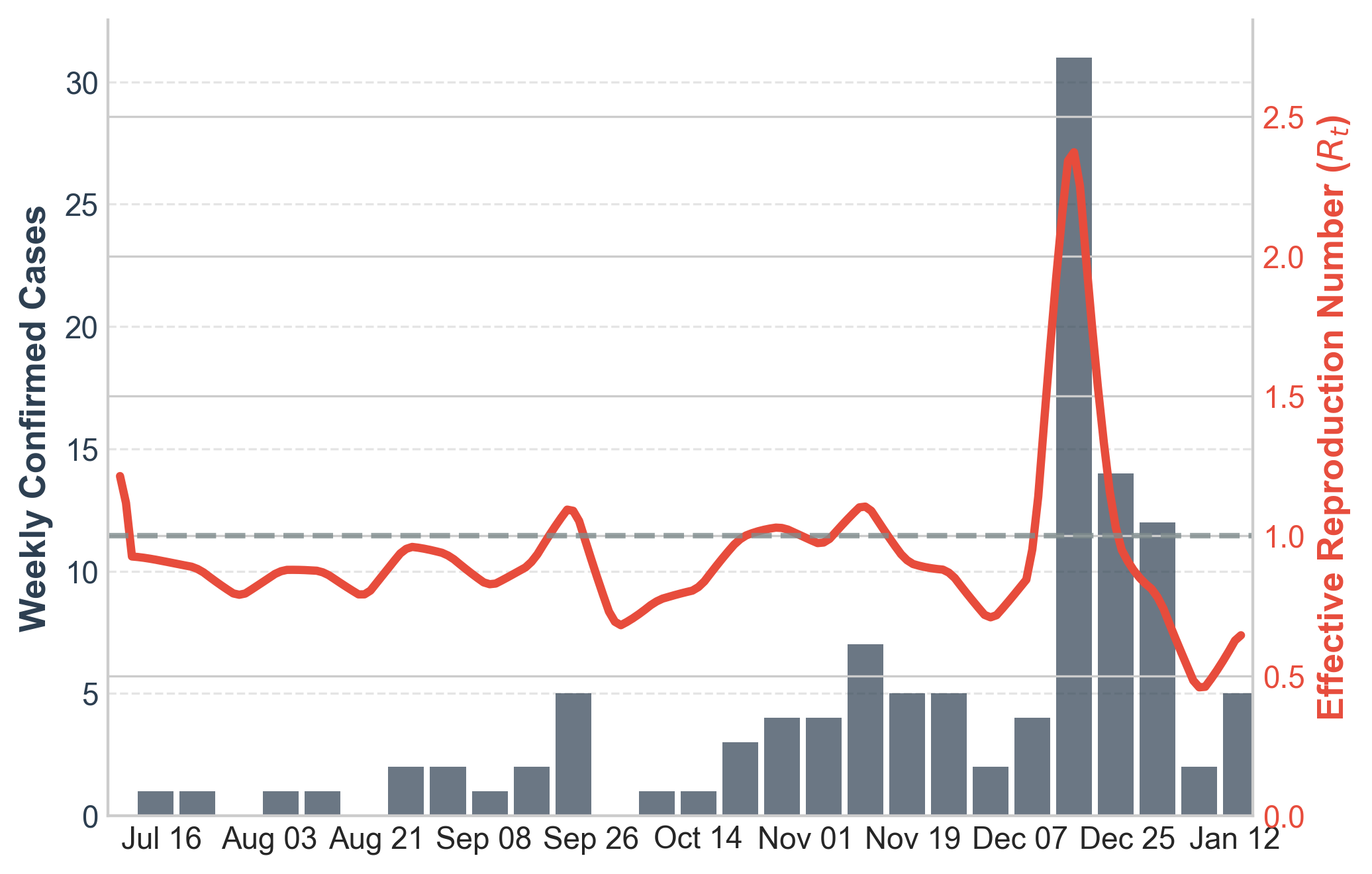

**Supplementary Figure S1. Historical Bundibugyo virus disease dynamics from the 2007 Bundibugyo outbreak.** Daily or reconstructed incidence and effective reproduction-number estimates used to calibrate the historical high-transmission robustness analysis.

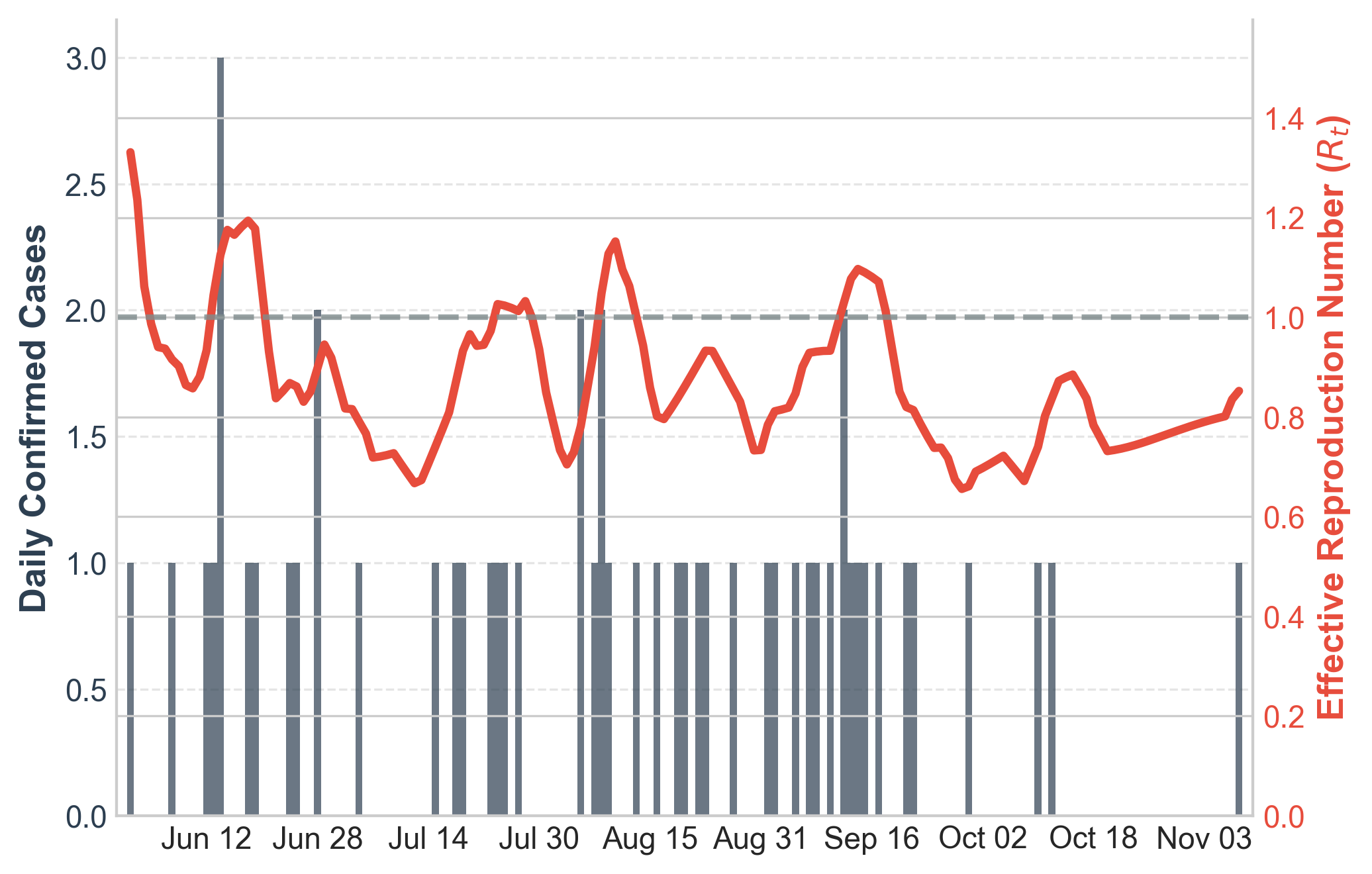

**Supplementary Figure S2. Historical Bundibugyo virus disease dynamics from the 2012 Isiro outbreak.** Daily or reconstructed incidence and effective reproduction-number estimates used to calibrate the second historical robustness analysis.

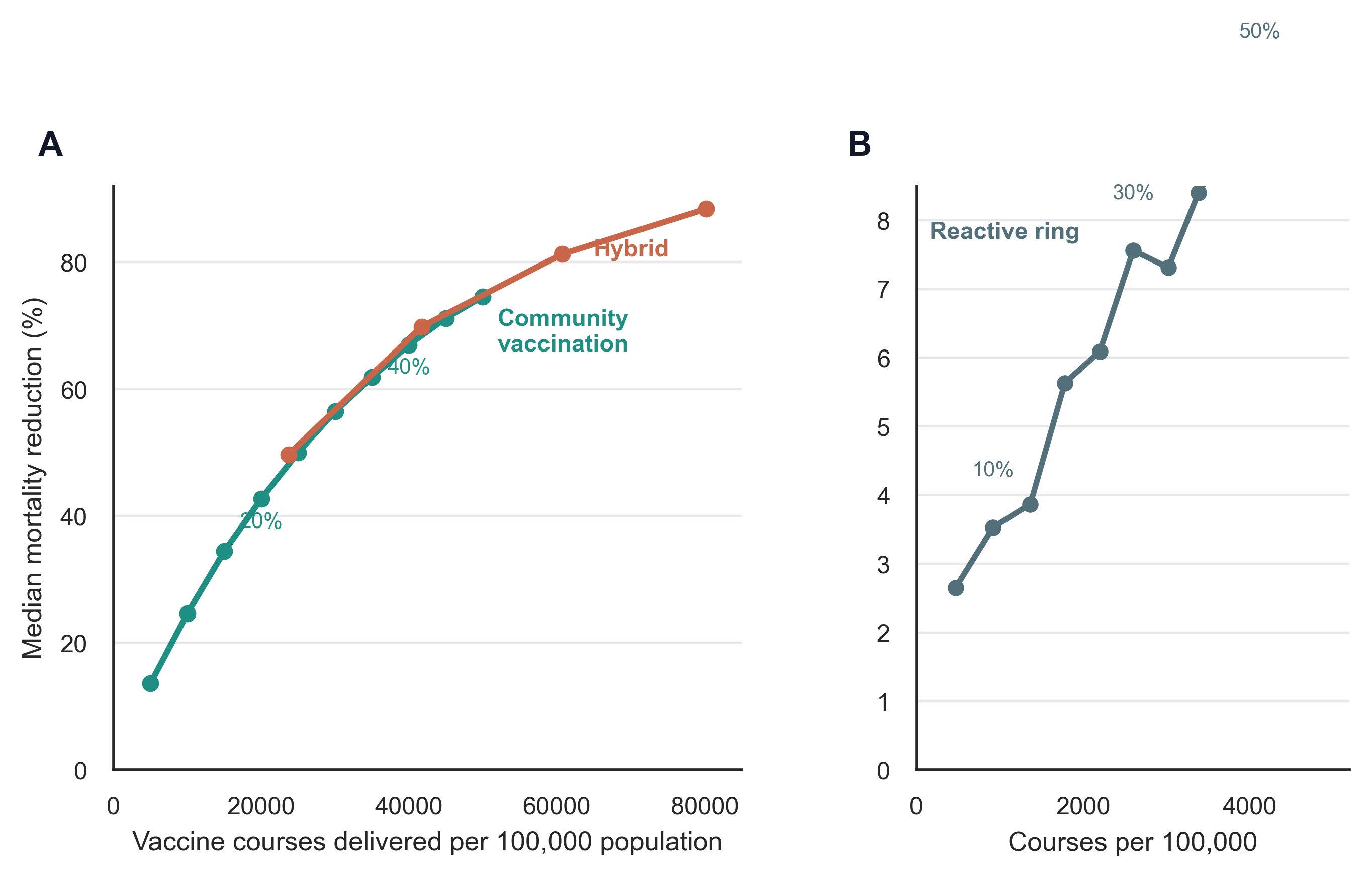

**Supplementary Figure S3. Dose-use frontier for reactive ring and community vaccination strategies.** Points show median mortality reduction and vaccine courses administered under the corrected 45% vaccine-effect assumptions. Comparisons are intended to show the trade-off between vaccine use and mortality reduction across operational strategies.

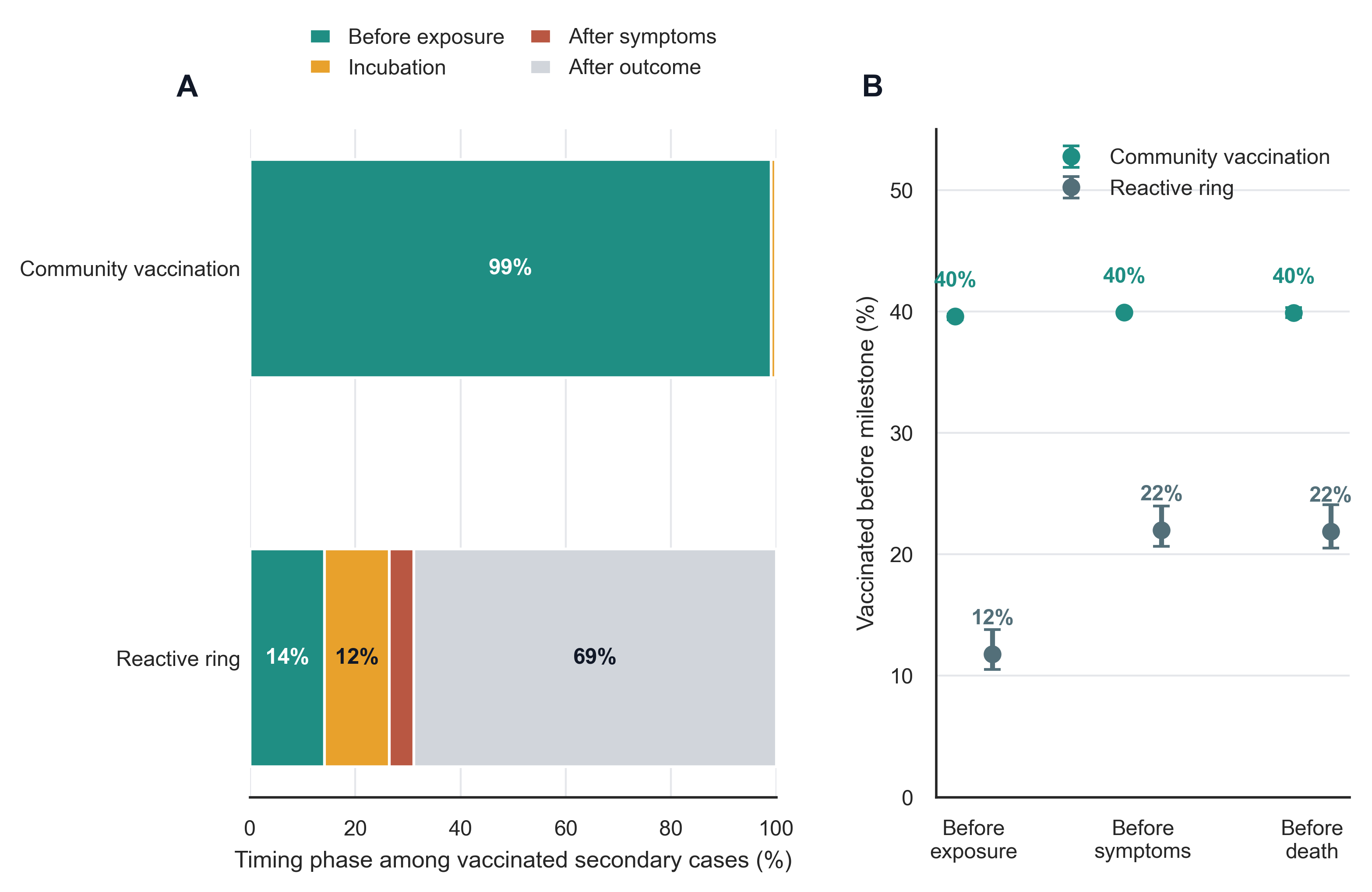

**Supplementary Figure S4. Delivery-window analysis.** Distributions show when vaccinated eventual infected individuals received vaccine relative to exposure and symptom onset under reactive ring and community vaccination strategies. Earlier vaccination relative to exposure is the main mechanism separating community-vaccination benefit from reactive ring vaccination
